# Chronic health conditions, multimorbidity and subsequent economic inactivity in three British birth cohorts

**DOI:** 10.64898/2026.11.02.26367051

**Authors:** Demelza Smeeth, Laura Gimeno, Charis Bridger Staatz, Jacques Wels, Nish Chaturvedi, George B Ploubidis

**Author notes:** Joint contributions (Demelza Smeeth, Laura Gimeno).

## Abstract

**Background:** Economic inactivity due to ill-health has been rising in the United Kingdom. Chronic health conditions make up most of the burden of poor health, and frequently co-occur, yet the impact of multimorbidity on economic inactivity remains underexplored.

**Methods:** Data was from three British birth cohorts (born in 1946, 1958 and 1970). Psychological distress, chronic back pain and cardiometabolic disease were assessed in participants’ thirties and forties, and economic inactivity one to two decades later. Modified Poisson regression models with robust standard errors estimated associations between health and inactivity, accounting for economic activity at baseline and several socioeconomic and demographic confounders. We tested for multiplicative and additive interaction between condition pairs.

**Results:** The prevalence of all three health conditions increased with age. Each was associated with a higher risk of later economic inactivity in fully adjusted models. Psychological distress was the strongest and most consistent predictor across cohorts (RR = 1.22 (95% CI 0.90-1.64) to 1.99 (95% CI 1.67-2.38)). Comorbidity was common; more than 10% of participants experienced two or more conditions in their forties. Risk of economic inactivity increased with multimorbidity. However, we found little evidence that risk for comorbidity exceeded the sum of individual conditions on either the multiplicative or additive scales.

**Conclusions:** Common chronic conditions in adulthood, particularly when comorbid, are associated with a substantially higher risk of economic inactivity across decades-long time horizons. This association has persisted over time and across policy contexts.

## Introduction

The number of working-age adults in the UK who are out of the labour market (economically inactive) because of long-term sickness has increased from 2 million in spring 2019, to over 2.8 million in Spring 2024.^1,2^ There is particular concern over rapidly rising rates of Not in Employment, Education or Training (NEET) among young people (aged 16-24).^3,4^ However, older age groups have also been affected with the greatest absolute increases in health-related inactivity observed among those aged 50-64.^5^ This trend is of particular concern because rising health-related economic inactivity among older adults has emerged as a prominent policy issue, with recent reports identifying it as a substantial constraint on labour-force participation, productivity, and economic growth in the UK.^1^ In the Labour Force Survey, the most frequently cited reasons for health-related inactivity are common and chronic in nature: mental health problems, musculoskeletal issues, and cardiometabolic conditions.^2,6–8^

Mental ill-health, musculoskeletal, and cardiometabolic conditions are not only highly prevalent, but also frequently co-occur.^2^ An estimated 14.8% of people in the UK have two or more long-term health conditions (multimorbidity), rising steeply with age,^9^ and becoming more common in more recently born generations.^6,10^ People with multimorbidity may be at greater risk of economic inactivity, because managing several conditions, often not affecting the same systems (e.g., mental health and physical health) can complicate disease management pathways.^11^ Understanding the extent to which individuals with multiple health conditions are at higher or lower risk of economic inactivity than those with single conditions, and whether this risk is greater than what could be predicted by each of the single conditions independently (i.e., presence of multiplicative or additive interaction) is important to inform targeted support.

Despite growing evidence of chronic multimorbidity as a population health challenge and clear concern over the impact of population health on the broader economy, little research to date has examined the impact of multimorbidity on work participation.^12,13^ Much of the existing research on the impact of ill-health on exit from work (including early retirement, all-cause labour market exit and unemployment) has explored the consequences of acute health shocks (e.g., heart attacks, accidents).^7^ Evidence on the impact of chronic conditions, which make up the greater burden of poor health in the UK is limited.^14^ Recent evidence on the link between chronic multimorbidity and economic activity is more limited still. Cross-sectional analyses indicate that employment rates are far lower among those with multiple health problems (ranging from 65% among those with 1 condition to just 29% among those with ≥5 conditions)^15^ but associations could be socioeconomically confounded or affected by reverse causality.

In this study, we address these gaps using data from three nationally representative British birth cohorts of people born in 1946, 1958 and 1970. We examine long-term associations between psychological distress, cardiometabolic disease, and back pain, and subsequent economic inactivity. We then assessed whether people experiencing more than one of these health conditions were at particularly high risk of economic inactivity, and whether the combined effects of the health conditions exceeded those expected from each condition individually.

## Methods

### Cohort

This study used data from three British birth cohort studies representative of people born in 1946, 1958 and 1970: the 1946 MRC National Survey of Health and Development (NSHD46; N=5,362 at birth),^16,17^ the 1958 National Child Development Study (NCDS58; N=17,415),^18^ and the 1970 British Cohort Study (BCS70; N=17,198).^19,20^ All three studies follow people born during one week of 1946, 1958 or 1970 in England, Wales and Scotland from birth to the present.

The studies have collected data on a wide range of topics across the life course, including physical and mental health and social circumstances. We primarily used questionnaire or interview data collected during cohort members’ thirties, forties, and fifties. All cohort participants provided written informed consent. Ethical approval for NSHD46 was obtained from the National Research Ethics Service Committee London (14/LO/1173), for NCDS58 and BCS70 from the NHS Multi-Centre Research Ethics Committee.

### Economic inactivity

The primary outcome was a binary indicator of economic activity status in participants’ early forties (NSHD46: 43y, NCDS58 and BCS70: 42y) and early fifties (NSHD46: 53y, NCDS58: 50y, BCS70: 51-54y). We grouped participants into economically inactive and economically active. Economic inactivity included people not working for any reason (e.g., poor health, disability, retirement) or who are unemployed (i.e., seeking employment). Economic activity included people currently working (full or part-time), in education, in training or looking after the home or family. Contrary to the classification used by ONS,^21^ we classified people who were looking after the home or family as “active” due to the significant number of women in NSHD46 who were looking after the home or family in their thirties due to employment norms at the time, but who eventually entered the workforce. A binary outcome was selected over greater stratification, due to people having multiple reasons for not working.

### Health problems

The primary exposures were binary classifications of the presence of psychological distress, cardiometabolic disease or back pain assessed in cohort members’ thirties (NSHD46: 36y, NCDS58: 33y and BCS70: 34y) or forties (NSHD46: 43y, NCDS58 and BCS70: 42y). Detailed information on exposure measurement can be found in the Supplementary Methods.

### Psychological distress

Psychological distress is a measure of poor mental health capturing anxiety and depressive symptoms measured using standard questionnaires.^22^ Clinically validated cut-offs were applied to the total score to identify cohort members with significant levels of psychological distress.^23^

### Cardiometabolic disease

Cardiometabolic disease is a cluster of interconnected chronic disorders including cardiovascular disease and metabolic dysfunction. Participants were classified as having cardiometabolic disease if they self-reported diagnosis of any cardiovascular complaints (angina, myocardial infarction, hypertension, etc.) or diabetes. For the measure in participants’ forties, we also carried forward the small number of cases identified in their thirties, under the assumption that these conditions are chronic and can be managed but are usually not cured.

### Back pain

Cohort members were classified as having back pain if they answered yes to a question about back problems, chronic or severe back pain, sciatica or lumbago (currently or in the last 12 months).

### Statistical analyses

We modelled the relationships between each health exposure and subsequent economic inactivity using modified Poisson regression with robust standard errors.^24^ Analyses were run separately for each cohort. We examined associations between health and economic inactivity over three timespans: health in thirties and inactivity in forties, health in forties and inactivity in fifties, and health in thirties and inactivity in fifties. For each model, we implemented three levels of covariate adjustment. Crude models included only health and inactivity. Minimally adjusted models additionally adjusted for economic activity assessed at the same time as the health condition (lagged outcome), age, and sex. The inclusion of the lagged outcome enabled us to model the move into economic inactivity. Fully adjusted models additionally controlled for a wide range of socioeconomic and demographic characteristics from across the life course (e.g., parental social class, household income, partnership status). More information on these covariates can be found in the Supplementary Methods.

Following the same procedure, we examined the role of multimorbidity. First, we ran models using the count of conditions as an exposure (0, 1, 2, or 3, with 0 as the reference category). We then assessed multiplicative and additive interactions between each possible pair of conditions. We tested for multiplicative interaction by adding an interaction term between pairs of conditions when both were included in the same model and tested for additive interaction using the Relative Excess Risk due to Interaction (RERI).

As secondary analysis, we used multinomial logistic regression models to examine the association between health and different types of economic inactivity, differentiating between those who were unemployed (out of work but looking for employment) and others. As sensitivity checks, we re-ran models classifying those looking after home or family as economically inactive and stratified by sex.

Like all longitudinal studies, the British birth cohorts are affected by attrition and by item-level missingness.^22^ For all analyses, we used multiple imputation with chained equations (MICE) to address both attrition and item non-response, by imputing missing data on all variables included in the models among the target population at the outcome sweep (forties or fifties), that is, all people who were alive and living in the UK at the time of data collection. Imputation was performed using the *mice* package creating twenty imputed datasets for each cohort.^25^ Since MICE works under the missing-at-random (MAR) assumption, we included auxiliary variables which are predictive of missingness and underlying missing values in our imputation models.^26,27^ Full details on the imputation procedure are outlined in the Supplementary Methods.

Analyses were conducted in R (v.4.5) running in RStudio.

## Results

### Poor health and economic inactivity throughout the working life

Across the three UK birth cohorts, the proportion of participants with psychological distress or cardiometabolic disease increased with age and was generally higher in more recent birth cohorts (*Figure 1*). Prevalences were lowest in NSHD46 at age 36 (psychological distress: 7.3%; cardiometabolic disease: 7.1%) and highest in BCS70 at age 43 (psychological distress: 22.6%; cardiometabolic disease: 16.7%). Back pain was also more common in participants’ forties in NSHD46 and BCS70, but the reverse was true for NCDS58 due to particularly high reported prevalence in their thirties.

**Figure 1.**
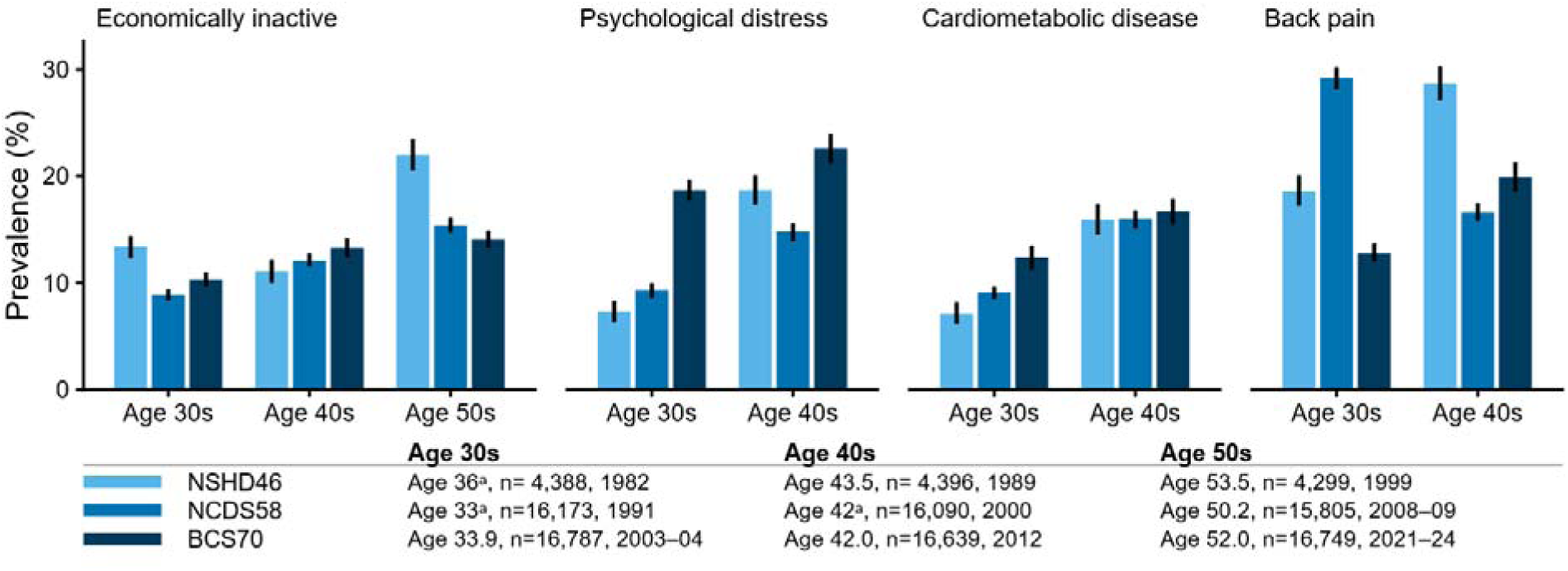
Cohort description. Depicts the prevalence of economic inactivity and each health condition across each cohort and decade combination alongside the year of data collection, sample size, and mean age of cohort members. Underlying values can be found in *Supplementary Table 2.* ^a^Age is only available as a round number in these sweeps.

Comorbidity was experienced by at least 4.6% of cohort members in either decade of life and became more common as participants aged, affecting at least 8.9% of cohort members in their forties (*Figure 2*). Psychological distress and back pain were the most frequently co-occurring conditions, reaching a prevalence as high as 4.7% in BCS70 at age 42. In their forties, at least 1% of participants in each cohort had all three conditions, though this level of comorbidity was rarer in their thirties.

**Figure 2.**
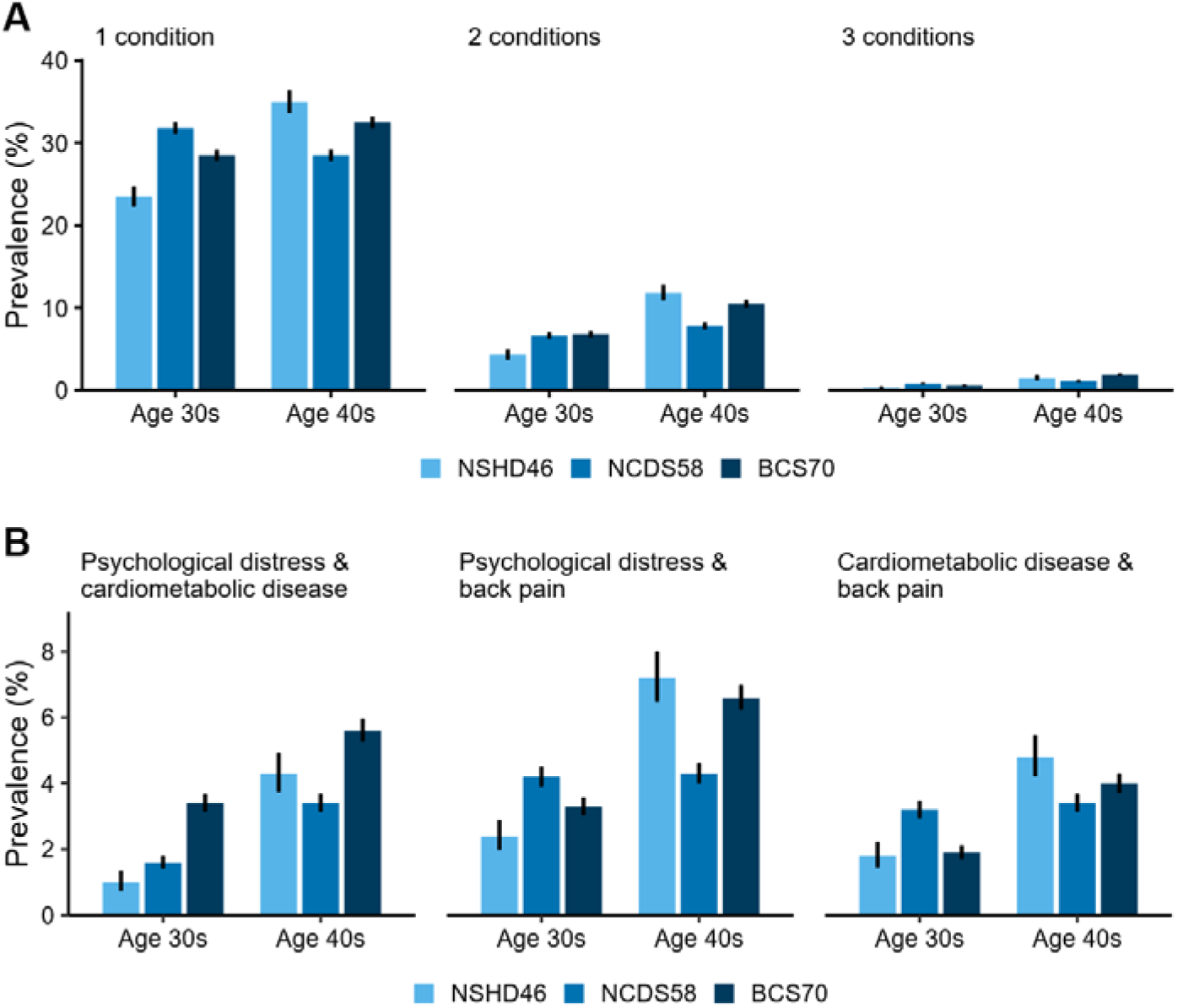
Prevalence of multimorbidity. A) The prevalence (%) of having one, two or 3 of the common health conditions. B) The prevalence of each specific pair of comorbid health conditions.

Economic inactivity, defined as those not in employment, education, training or caring roles, was experienced by 8.9-13.4% of cohort members in their thirties and forties (*Figure 1, Supplementary Table 2*). The prevalence of economic inactivity increased with age in all cohorts, with the highest rate observed for in NSHD46 at age 53 (22%). When cohort members were in their thirties, those seeking employment made up approximately half of the economic inactivity group, but at older ages, particularly by their fifties, most cohort members who were inactive were not actively seeking employment (*Supplementary Table 3*).

### Impact of poor health on economic inactivity

Cardiometabolic disease and psychological distress were consistently associated with a higher risk of becoming economically inactive one or two decades later in all three cohorts (*Figure 3, Supplementary Figure 1*). Fully adjusted risk ratios (RR) ranged from 1.31 (95% CI 0.90-1.22) to 1.85 (95% CI 1.55-2.20) for cardiometabolic disease, and from 1.22 (95% CI 0.90-1.64) to 1.99 (95% CI 1.67-2.38) for psychological distress. Back pain was also associated with economic inactivity in fully adjusted models, except when examining economic inactivity at age 43 in NSHD46, with RRs ranging from 0.95 (95% CI 0.75-1.21) to 1.43 (95% CI 1.24-1.65). Point estimates from the fully adjusted models were smaller than in the crude and minimally adjusted models (*Supplementary Figure 2*). Results were similar when caring responsibilities were redefined as economic inactivity (*Supplementary Figure 3*) and sex-stratified analyses revealed minimal sex differences (*Supplementary Figure 4*).

**Figure 3.**
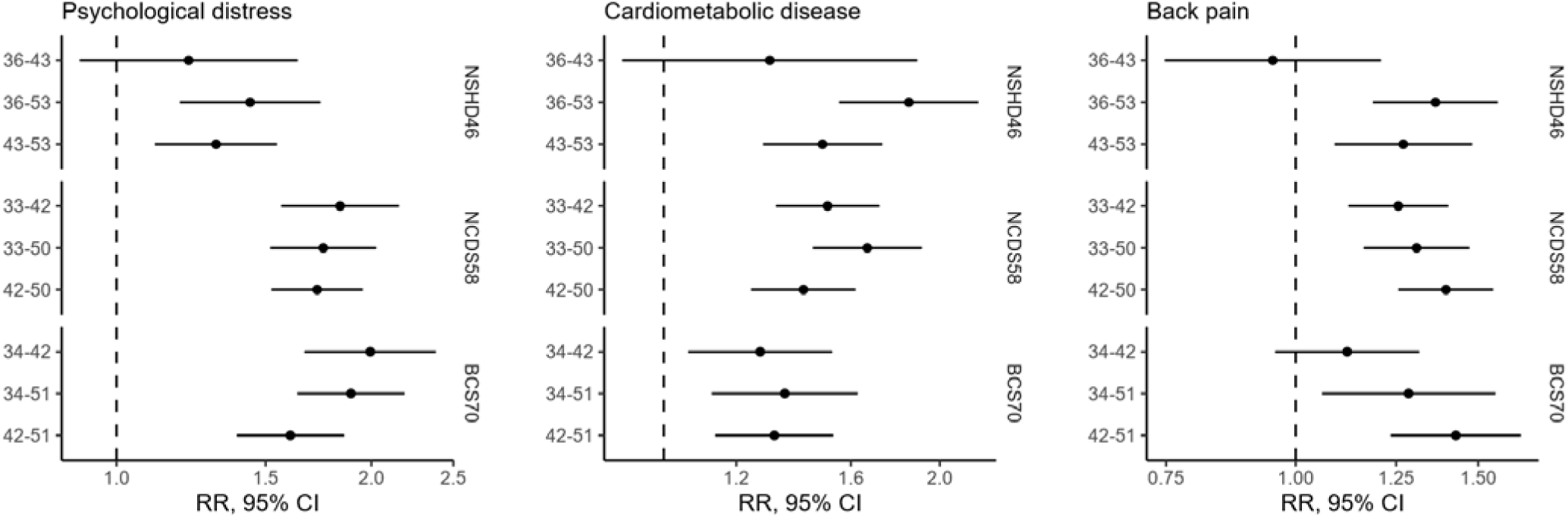
The association between health conditions and later transition to economic inactivity across the UK birth cohorts. Age pairings represent age at health assessment and economic activity assessment. Statistics derived from fully adjusted models. RR: risk ratio, CI: confidence intervals.

When we considered those inactive and looking for work separately from those inactive and not looking for work, we found that ill-health was more strongly associated the latter (*Supplementary Figure 5)*. However, despite weaker associations, in several cases, poor health was still significantly associated with unemployment, particularly for psychological distress.

### Impact of multimorbidity on economic inactivity

Those who had a single health condition consistently had a greater risk of subsequently becoming economically inactive than those without any health condition (1.28 (95% CI 0.89-1.82) to 1.77 (95% CI 1.58-1.98)). However, the risk for those with more than one health condition was greater still (*Figure 4*). This was observed for those with either two (1.65 (95% CI 1.39-1.97) to 3.35 (95% CI 2.83-3.97)) or three of the health conditions (1.71 (95% CI 0.35-8.25) to 5.34 (95% CI 4.19-6.79)). Associations were strongest for multimorbidity in participants’ forties, particularly in the more recent cohorts.

**Figure 4.**
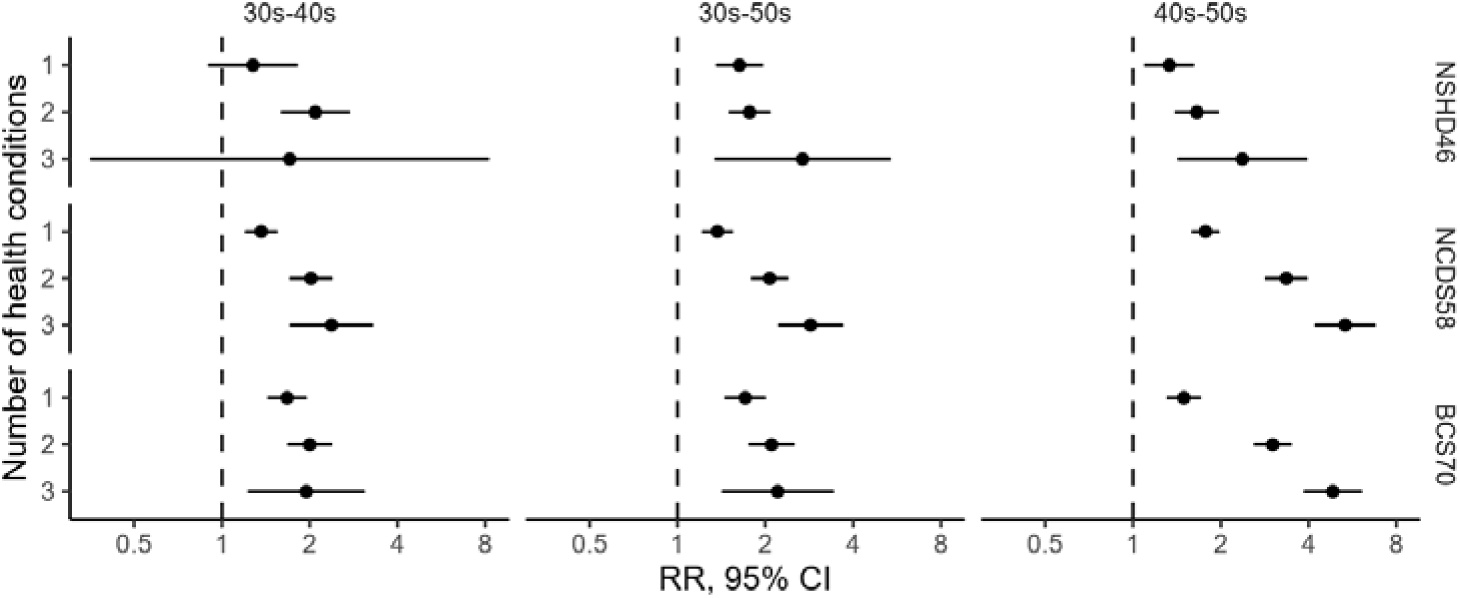
The association between number of health conditions and later transition to economic inactivity across the UK birth cohorts. Statistics derived from fully adjusted models. RR: risk ratio, CI: confidence intervals.

*Figure 5* shows the risk of becoming economically inactive for participants exposed to each of three pairs of health conditions (black dot, 95% CI), the excess risk attributable to each exposure alone (coloured bars) and the departure from additivity (RERI; hatched area), which is super-additive when it extends above the bars and sub-additive when it overlaps them. In most cases, each condition alone was associated with a higher risk of becoming economically inactive than participants with neither condition, and the point estimate was highest among participants with both conditions (*Supplementary Table 4*). Compared with participants with neither condition, risk ratios for psychological distress and cardiometabolic disease were generally larger than those for back pain (RR: back pain: 1.03 to 1.44; psychological distress 1.21 to 2.08; cardiometabolic disease 1.14 to 1.85). Consequently, when comorbid with back pain, psychological distress and cardiometabolic disease tended to account for the greater share of the excess risk of economic inactivity. Reflecting this, joint risk ratios were generally highest for comorbidity of psychological distress and cardiometabolic disease (RR: 1.61 (95% CI 0.68-2.54) to 2.53 (95% CI 1.90-3.17)).

**Figure 5.**
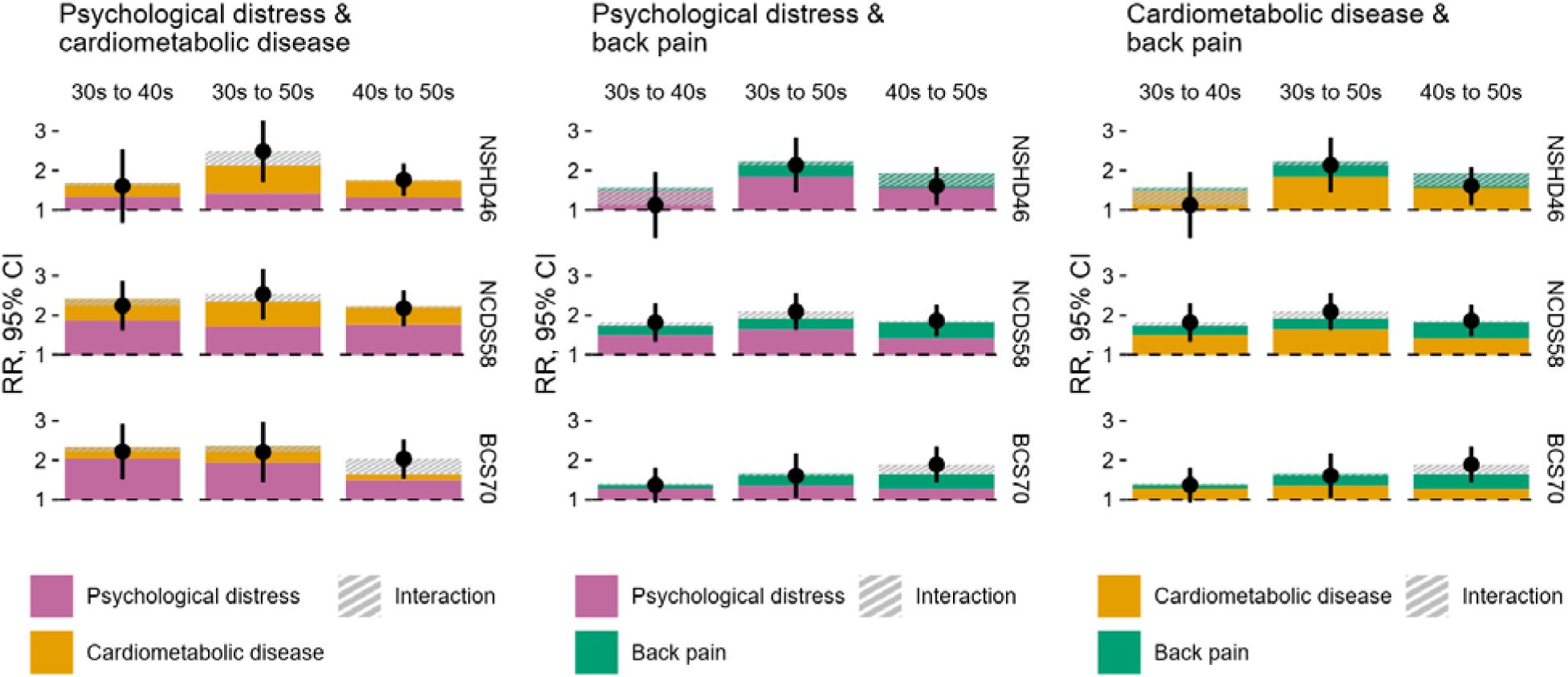
The association between pairs of health conditions and later transition to economic inactivity across the UK birth cohorts. Black points and error bars indicate the estimated RR (95% CI) for individuals exposed to both factors relative to the unexposed reference group. Stacked coloured bars decompose the joint association into the excess risk associated with each exposure individually and the additive interaction component, measured as the relative excess risk due to interaction (RERI). A positive RERI indicates that the joint effect exceeds that expected under additivity, whereas a negative RERI indicates that the joint effect is less than expected under additivity. Statistics derived from fully adjusted models. RR: risk ratio, CI: confidence intervals.

There was little evidence that effect of comorbidity on inactivity significantly exceeded (or fell below) the sum of the individual effects of the health condition pairs. On the additive scale, interaction coefficients varied across cohorts, comorbid pairs and ages, and confidence intervals included the null (RERIs: -0.55 (95% CI -1.21,0.10) to 0.46 (95% CI -0.81,1.72)). On the multiplicative scale, interactions tended to be more consistently below one (0.64 (95% CI 0.42,0.98) to 1.26 (95% CI 0.94,1.67) suggesting some degree of effect antagonism, however confidence intervals also included the null. Small numbers of events precluded our ability to explore the interactions between all three health conditions.

## Discussion

In the UK, working age adults with cardiometabolic disease, back pain or psychological distress were up to twice as likely to be economically inactive ten or twenty years later, even after accounting for previous employment status and a diverse set of confounders (though effect sizes varied by condition, cohort and age). These associations were observed despite the substantial changes in economic and policy contexts that happened between the early 1980s (when NSHD46 were in their early forties) and the 2020s (when BCS70 reached their early fifties). Our work extends beyond the findings of cross-sectional studies, which have shown that people who are not working due to health state mental ill-health, cardiometabolic disease and musculoskeletal pain as the reason for their inactivity.^15^ We find using prospectively collected data that ill-health has repercussions for economic inactivity, increasing the likelihood that these relationships are not a consequence of respondents attributing their labour market status to health problems. We also find that that ill-health has repercussions for economic inactivity regardless of whether participants identify health as the primary reason for not working.

Poor mental health has received considerable attention because of its association with economic inactivity at younger ages.^2^ We found that psychological distress was the condition most strongly and consistently associated with subsequent inactivity in all three cohorts. Although concerns about the economic consequences of poor mental health have intensified in recent years, our results indicate that the link between poor mental health and economic inactivity is not a recent phenomenon. It reflects a longstanding association that may be becoming increasingly important as mental health problems become more common.^28^ The high prevalence of poor mental health, together with its strong effect on health-related economic inactivity, identifies it as a key target for prevention and intervention.

In all three cohorts, nearly 1-in-10 cohort members had two or more of the conditions concurrently in their mid-forties. Comorbidity was associated with a greater risk of subsequent economic inactivity. Compared to those with no conditions, those with two conditions were up to 4 times more likely to become economically inactive, and those with three conditions were up to 7 times more likely. However, we found little evidence that comorbidity was associated with a greater risk beyond what would be expected from the contributions of the individual conditions, either on the additive or multiplicative scale. This should not be interpreted as evidence that comorbidity is unimportant. Rather, our results suggest the elevated risk associated with multiple conditions may reflect the accumulation of independent health burdens, which together substantially increase the likelihood of labour market exit. These findings highlight the importance of addressing health conditions individually, focusing on those which are the most strongly associated with economic inactivity such as cardiometabolic health or psychological distress. This is particularly relevant given that effective interventions are likely to differ across physical and mental health conditions, requiring coordinated and holistic approaches to support labour market participation. However, it should be noted that our definition of multimorbidity is conservative, only encompassing three categories of health conditions and combining related but distinct conditions such as hypertension and diabetes. Therefore, we may be underestimating the true association between multimorbidity and economic inactivity.

We found that ill health is sometimes associated with a higher risk of unemployment (not working but still looking for work), although usually less strongly than for those who are inactive and not looking for work. Because unemployed people are typically classified as economically active (e.g., by the Office for National Statistics),^21^ the importance of health may be overlooked as a risk factor for unemployment. Preventing ill-health and better supporting people experiencing chronic health problems may therefore also have benefits for reducing unemployment, yet current interventions to reduce unemployment tend not to focus on this area, instead emphasising training, assistance, and sanctions.^29^

A major strength of our study is the exceptionally rich longitudinal data offered by the British birth cohort studies. Data collection has spanned decades in three generations, and this enabled us to explore long-term repercussions of chronic health conditions on subsequent economic activity across different policy, economic and social landscapes. Since chronic health conditions may not immediately affect a person’s ability to remain in employment, studies with shorter follow-up time may substantially underestimate the full economic burden associated with poor health. As data on many aspects of life has been collected prospectively since birth, we were able to tightly control for factors associated with both health and labour market outcomes, strengthening causal inference in an area where experimental studies are not possible.

Heath conditions were self-reported in our study, which could be seen as a weakness. However, this also means that our exposure measures do not just capture those who have been in contact with health services and received a formal diagnosis. This is important because mental ill-health and back pain may affect labour market participation but are frequently unreported to the healthcare system. This means that the self-reports used here likely better reflect the true population burden of these conditions, capturing individuals who would otherwise be missed. This is likely especially true for the NSHD46, for whom the diagnostic criteria, awareness, and stigma surrounding mental health diagnoses has changed dramatically over the course of their lives.

This study also has limitations. As with all longitudinal studies, the birth cohorts have suffered from attrition, however, the extensive information collected on participants throughout life and the known properties of the sample at baseline meant that we were able to use multiple imputation to restore representativeness to the target population. The birth cohorts are also largely ethnically homogeneous (>95% white), which precluded our ability to explore ethnic differences in health-inactivity associations. Due to limited power, we were also unable to further disaggregate analyses by demographic and socioeconomic characteristics beyond sex. Future work, using larger-scale administrative data, could explore whether and how the health-work associations vary by population subgroup. Finally, differences in health measurement across cohorts limited our ability to identify secular changes in the observed relationships, but there was little suggestion that associations had weakened across cohorts, consistent with findings from previous research comparing NCDS58 and BCS70.^14^

Over 1 in 5 working-age adults in the UK are currently out of work and not seeking work.^1^ Our findings show that common chronic health conditions in adulthood have repercussions for economic inactivity that can be observed a decade or so later, with the greatest risk among individuals living with multimorbidity. The prevalence of several health conditions, particularly mental ill-health and multimorbidity, is higher in more recent cohorts compared to predecessors at the same age.^6^ Our findings therefore raise concerns about future workforce participation and the sustainability of labour market engagement without intervention.^6^ Addressing this challenge will require coordinated action across health, employment, and social policy. Although the UK Government has increasingly recognised the interdependence of health and work,^1^ effective interventions will need to account for the complexity and heterogeneity of health conditions and individual socioeconomic circumstances that shape people’s risk of labour market exit.

## Data Availability

NSHD data used in this publication are available to bona fide researchers upon request to the NSHD Data Sharing Committee via a standard application procedure. Further details can be found at http://www.nshd.mrc.ac.uk/data; doi: 10.5522/NSHD/Q101; doi: 10.5522/NSHD/Q102. NCDS and BCS70 data used in these analyses can be downloaded by registered users via the UK Data Service, free of charge (https://ukdataservice.ac.uk/).

## Acknowledgments

We are extremely grateful to the NSHD, NCDS and BCS70 study members for their lifelong participation and continuing support; and to past and present members of the study teams, who helped to collect and process the data. NSHD data used in this publication are available to bona fide researchers upon request to the NSHD Data Sharing Committee via a standard application procedure. Further details can be found at http://www.nshd.mrc.ac.uk/data; doi: 10.5522/NSHD/Q101; doi: 10.5522/NSHD/Q102. NCDS and BCS70 data used in these analyses can be downloaded by registered users via the UK Data Service, free of charge (https://ukdataservice.ac.uk/).

