## Supplemental information for "Chronic health conditions, multimorbidity and subsequent economic inactivity in three British birth cohorts"

### Supplementary Methods

#### Health problems

The exposures were binary classifications of the presence of psychological distress, cardiometabolic disease or back pain assessed in cohort members’ thirties (NSHD46: 36y, NCDS58: 33y and BCS70: 34y) or forties (NSHD46: 43y, NCDS58 and BCS70: 42y).

##### Psychological distress

Psychological distress is a measure of poor mental health capturing anxiety and depressive symptoms,^1^ and can be measured using standard questionnaires. This measure of mental health was chosen due to the significant proportion of cases where individuals do not seek medical attention, diagnosis or treatment, and how these have changed over subsequent generations: using a measure based on diagnosed mental health conditions alone would risk severely underestimating the presence of mental ill-health, particularly in the oldest generations. This can be observed by the small number of individuals who take antidepressants (1.3-2.3%) despite the relatively high prevalence of psychological distress (7-19%) in NSHD46. In NSHD46, mental health was assessed in the cohorts using the Present State Examination at 36y and Psychiatric Symptom Frequency Scale at 43y. In NCDS58 and BCS70, it was measured using the 9-item Malaise Inventory. Clinically validated cut-offs were applied to the total score to identify cohort members with significant levels of psychological distress (PSE ≥ 5, PSFS ≥ 23, or Malaise ≥ 4).^2^

##### Cardiometabolic disease

Cardiometabolic disease is a cluster of interconnected chronic disorders including cardiovascular disease and metabolic dysfunction. Participants were classified as having cardiometabolic disease if they self-reported a doctor’s diagnosis of any cardiovascular complaints (angina, myocardial infarction, stroke, hypertension, etc.) or diabetes at that timepoint. For the measure in participants forties, we carried forward any reports of these conditions in their thirties to capture lifetime prevalence of cardiometabolic disease, under the assumption that these conditions are chronic rather than intermittent. In all three cohorts, and particularly in NCDS58 and BCS70, the great majority of people identified as having cardiometabolic disease have high blood pressure or diabetes.

##### Back pain

Cohort members were classified as having back pain if they answered “yes” to a question about currently having back problems, chronic or severe back pain, sciatica or lumbago (or having experienced these problems within the last 12 months).

#### Covariates

Time-invariant covariates were measured in early life. Sex was recorded at birth and age at interview was calculated using birth year and time of interview. Early life socioeconomic status was based on their father’s self-reported occupation at age 10 or 11 and categorised as manual or non-manual according to the Registrar General Social Class classification (non-manual: RGSC I, II and III-NM; manual: RGSC III-M, IV and V) ^3^. Childhood cognitive ability was assessed at age 10 or 11. Our measure is a composite score based on performance across a battery of cognitive tests covering verbal, non-verbal, numerical skills. Educational attainment was categorised as achieving a qualification equivalent to National Vocational Qualification (NVQ) level 4 or above.

Model-specific covariates were assessed at the same time as health. Partnership status was self-reported and measured using a binary variable indicating marriage or the presence of a cohabiting long-term partner. Household income was self-reported. Housing tenure was self-reported and measured using as a binary variable representing whether the participant owned their own home (fully or with a mortgage). Household size was self-reported and measured using as a binary variable representing more than two people living in the household (NSHD46) or the presence of children (biological or non-biological) living in the household (NCDS58 and BCS70).

#### Imputation

Like all longitudinal studies, NSHD46, NCDS58 and BCS70 have suffered from attrition over time. The likelihood of non-response is patterned by socioeconomic characteristics. For instance, individuals who are male, with lower socioeconomic status, and less healthy (obesity, lower self-rated health, smoking), are more likely to drop out from the study.^4–6^

Because of the richness of the data collected by the birth cohort studies across the life course and a wide range of domains, and the known properties of the cohorts at baseline, it is possible to take principled approaches to missing data to address biases introduced by item nonresponse (i.e., not answering a particular question despite responding to a particular survey) and attrition (i.e., not answering a particular survey). Methods such as multiple imputation and inverse probability weighting for non-response have been shown to successfully restore the representativeness of the birth cohort to the initial sample and to external benchmarks.^4,5,7–9^

We used multiple imputation by chained equations (MICE) to mitigate the impact of both item non-response and attrition. We define the target population for each model as cohort members who were alive and living in Britain at the time of the outcome sweep. Individuals who are not in the target population are those who had died by the time of the outcome sweep, or who had emigrated.

MICE was used to impute missing data on the exposures, confounders and the outcome among the target population, using the mice package in R.^10^ MICE functions under the Missing At Random (MAR) assumption, that is, the assumption that missingness is random conditional on observed variables.^11^ While this assumption is largely untestable, it can be made more plausible by including a rich set of auxiliary variables which are predictive of nonresponse and the underlying missing values themselves.^5^ A list of the auxiliary variables used in each imputation model are shown in Table S1 below. We conducted imputation for each cohort and analytical age combination, generating 20 imputed datasets each time. Model estimates were combined across imputations following Rubin’s rules.

Since an important component of the analyses in this paper revolved around understanding multimorbidity, it was important to allow for interactions between health conditions in the imputation models themselves. Failure to do so could bias any interaction test towards the null. To do this, we followed the approach used by Tilling and colleagues and included the interaction terms in the imputation models for all model variables.^12^

#### Modelling

We used modified Poisson models with robust standard errors to examine the relationship between health status and subsequent economic inactivity. This approach has the advantage of producing Risk Ratios (RR) which are easier to interpret than Odds Ratios (ORs), particularly when the outcome is common, and overcomes issues of non-collapsibility of the OR.^13,14^

For each exposure-outcome combination within each cohort, we created three models, accounting for different levels of confounding. Crude models only included the health indicators and economic inactivity at the outcome sweep.  Minimally adjusted models additionally controlled for a lagged economic inactivity term assessed at the same time as the health condition(s), to model the move into that economic inactivity group, as well as age at outcome measurement and sex. Fully adjusted models additionally adjusted for parental socioeconomic status (age 10/11), cognitive ability in childhood (age 10/11), and educational attainment, housing tenure, partnership status, household income, and number of children or total people in the household at the time of health assessment.

Confounders were selected according to *a priori* knowledge about their relationship to health status and economic outcomes. The hypothesised relationships between variables included in the analysis are shown in the Directed Acyclic Graph (DAG) below (Figure S1). In addition to the set of other demographic and socioeconomic controls, the inclusion of the lagged outcome helps to increase the plausibility that our analyses reflect the impact of ill-health on economic activity, by blocking confounding pathways from potential unmeasured confounders.

### Supplementary Figures

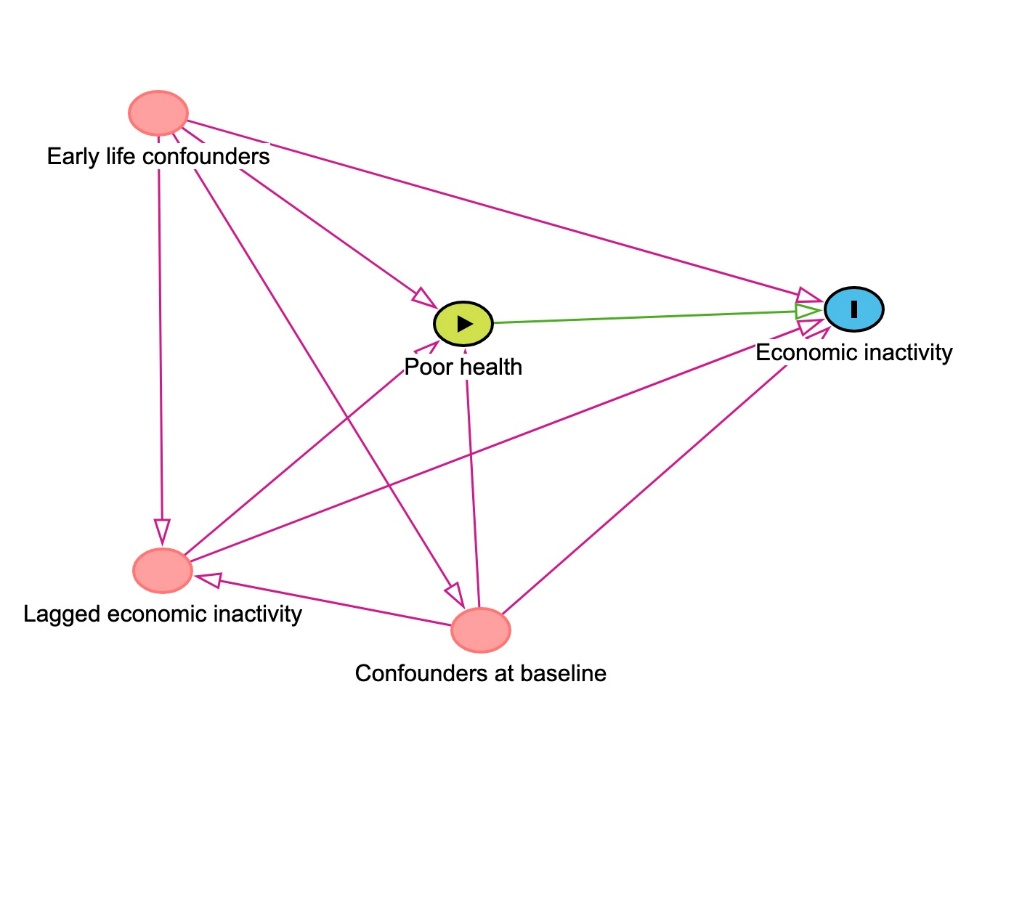

**Supplementary Figure 1.** **Directed acyclic graph (DAG)** **outlining the assumed causal relationships between health, confounders, and economic activity.**

Produced using DAGitty.^15^

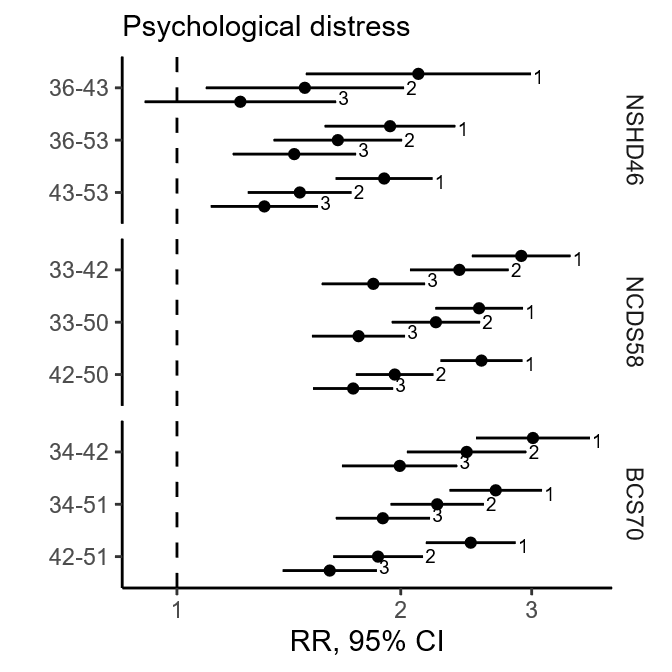

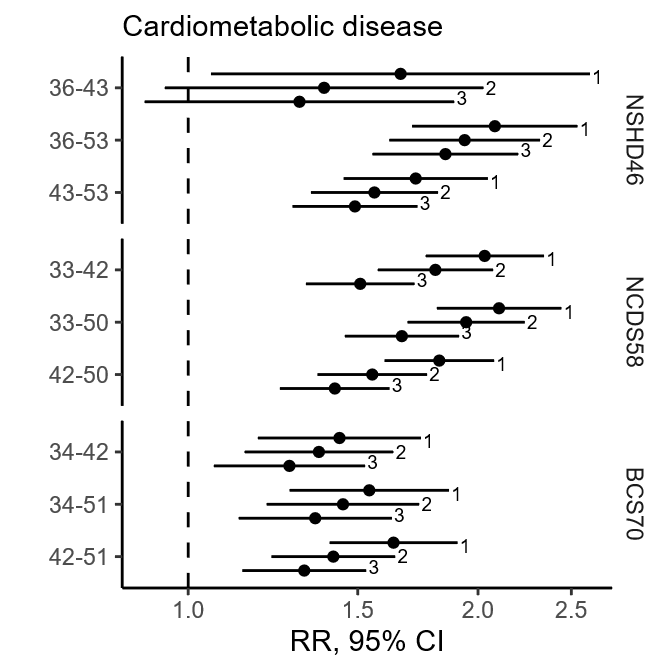

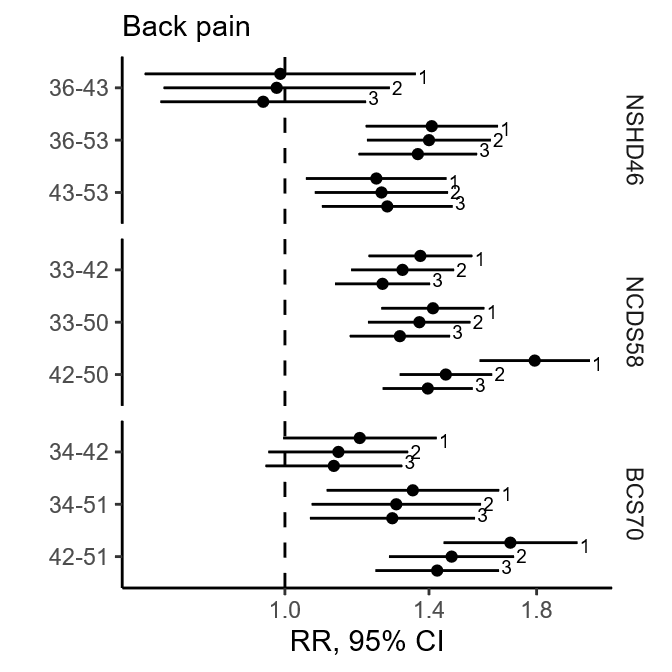

**Supplementary Figure 2. The association between health conditions and later transition to economic inactivity across the UK birth cohorts.**

Age pairings represent age at health assessment and economic activity assessment. Bars numbered according to level of adjustment. 1: crude models, 2: minimally adjusted, 3: fully adjusted. RR: risk ratio, CI: confidence intervals.

**Supplementary Figure 3. The association between health conditions and later transition to economic inactivity across the UK birth cohorts when caring was categorised as economic inactivity**

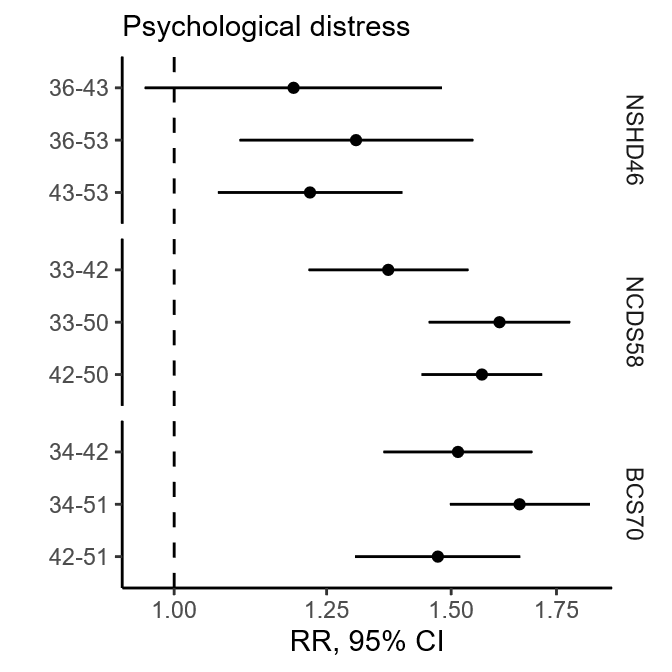

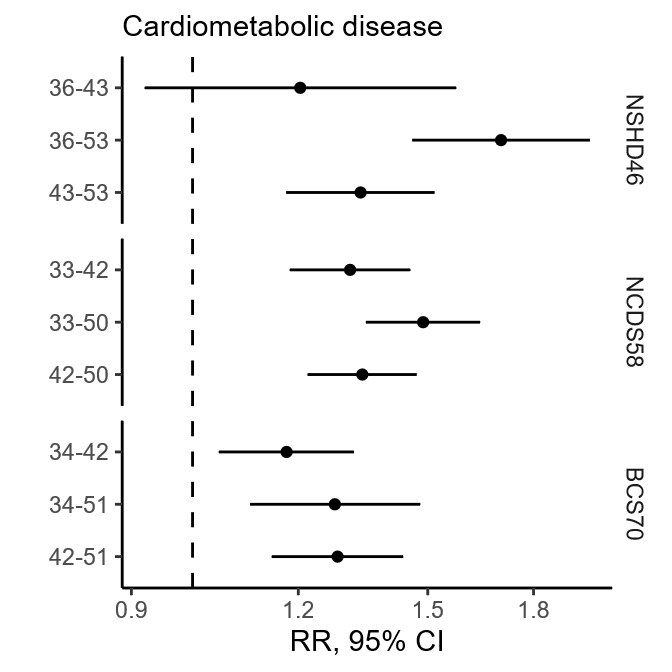

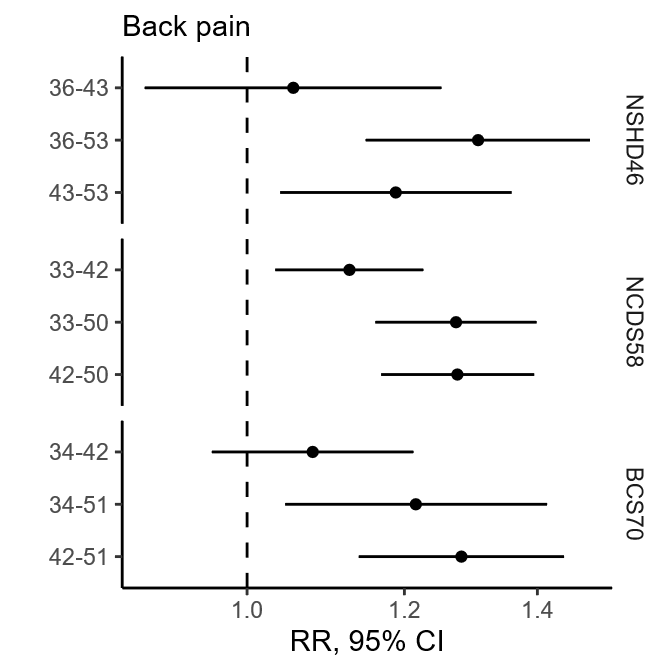

Age pairings represent age at health assessment and economic activity assessment. Statistics derived from fully adjusted models. RR: risk ratio, CI: confidence intervals.

**Supplementary Figure 4. Sex-stratified associations between health conditions and later transition to economic inactivity across the UK birth cohorts**

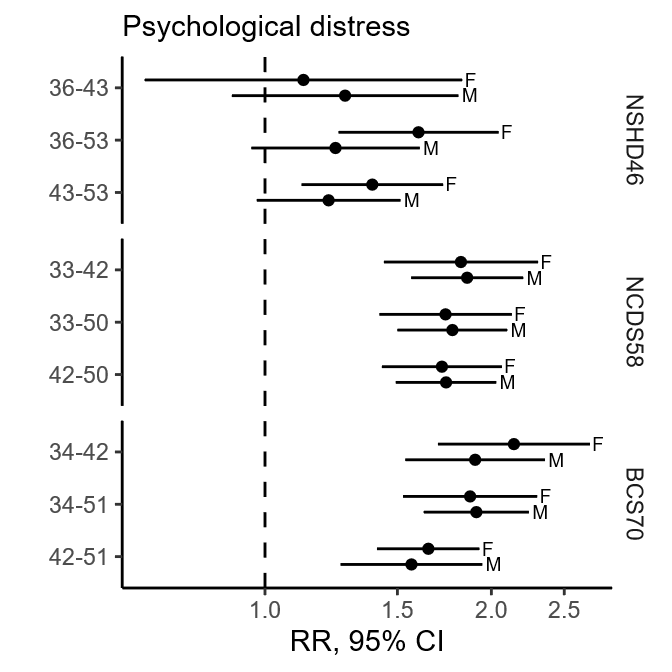

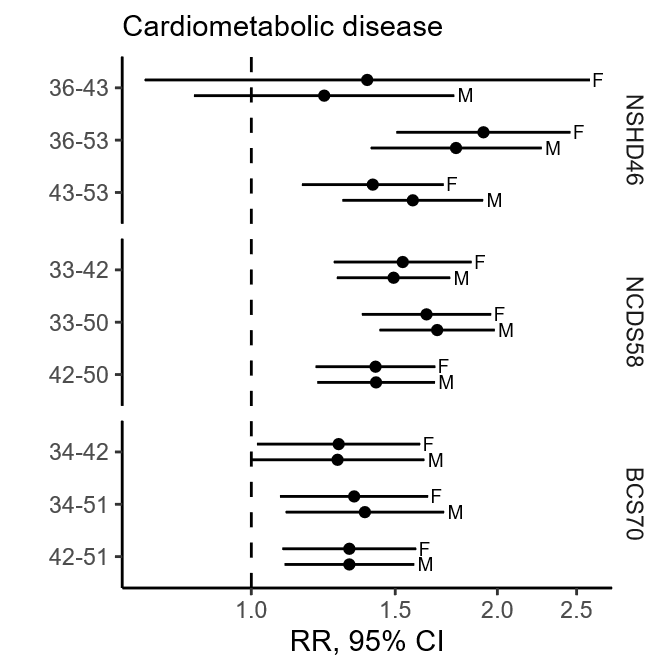

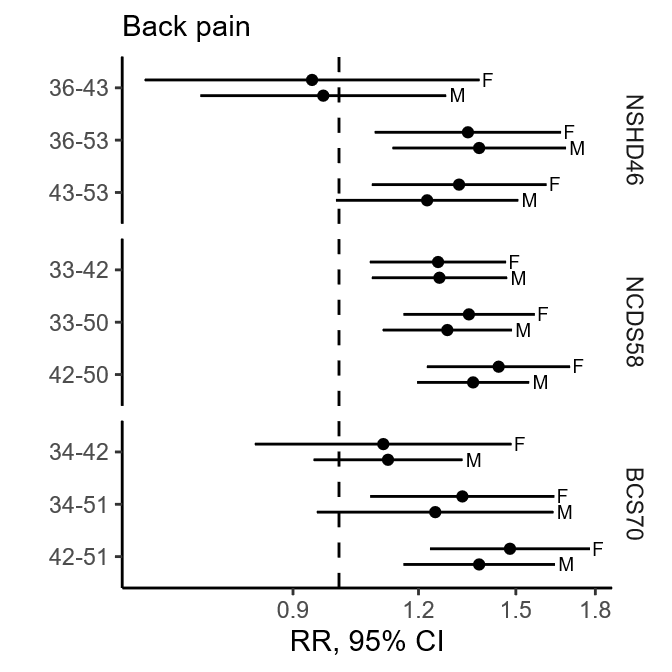

Age pairings represent age at health assessment and economic activity assessment. Statistics derived from fully adjusted models. RR: risk ratio, CI: confidence intervals.

**Supplementary Figure 5. Associations between health conditions and later transition to economic inactivity according to whether they were looking for work.**

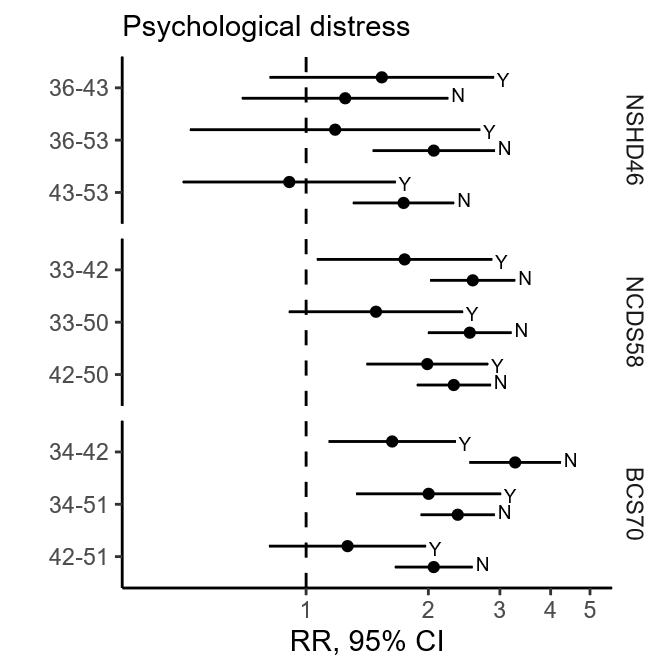

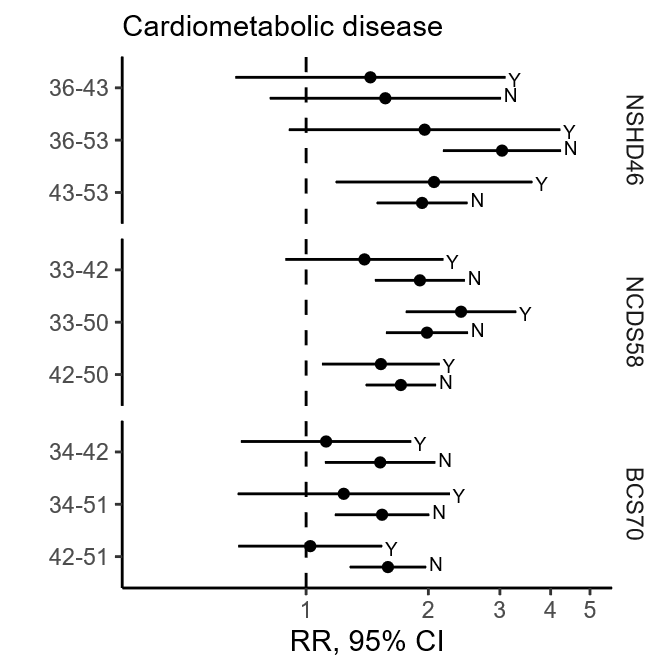

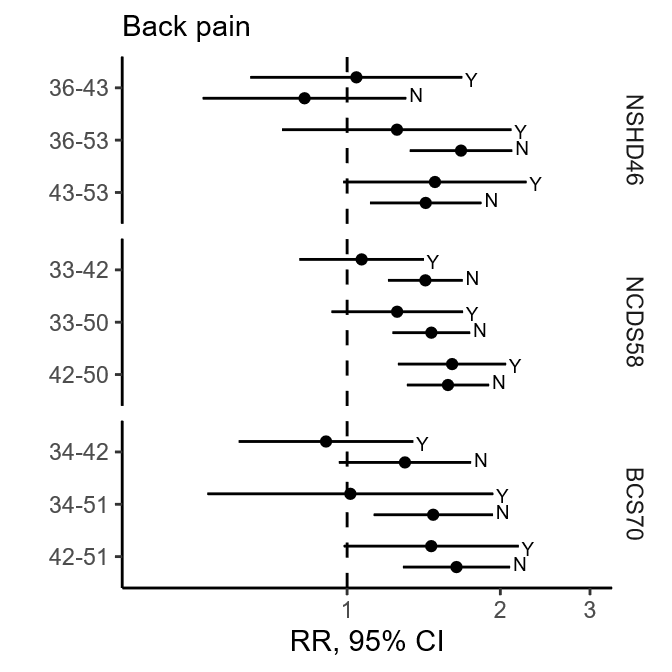

Age pairings represent age at health assessment and economic activity assessment. Statistics derived from fully adjusted models. RR: risk ratio, CI: confidence intervals.

### Supplementary Tables

| Supplementary Table 1. Imputation auxiliary variables | | | |
| --- | --- | --- | --- |
|  | **NSHD46**  **Age (years)** | **NCDS58**  **Age (years)** | **BCS70**  **Age (years)** |
| ***Early life*** |  |  |  |
| Childhood mental health | 13 | 11 | 10 |
| Adolescent mental health | 20 | 16 | 16 |
| Crowding | 0-11 | 0 | 0 |
| Birthweight | 0 | 0 | 0 |
| Childhood BMI | 11 | 11 | 10 |
| Adolescent BMI | - | 16 | 16 |
| Maternal education | 6 | 0 | 0 |
| Paternal education | 6 | 0 | 0 |
| Country of birth | 0 | 0 | 0 |
| Chronic health conditions | 5 | 11 | 10 |
| Childhood disability | 13 | - | - |
| Ever breastfed | 2 | 7 | 5 |
| Housing tenure | 4 | 7 | 5 |
| Maternal smoking | - | 0 | 0 |
| Sole access to amenities | - | 11 | - |
| ***Adulthood*** |  |  |  |
| Longstanding illness | 53 | 33, 43 | 34, 42 |
| Age left full-time education | 26 | - | - |
| Smoking | 36, 43, 53 | 33 | 34 |
| Voting | 26 | 33 | 34 |
| Member in organisations/unions | 26 | 33 | 34 |
| Self-rated health | - | - | 34 |
| Adult BMI | 36, 43, 53 | 33, 42 | 34, 42 |
| Number of previous sweeps | 0-64 | 0-34 | 0-34 |

| Supplementary Table 2. Additional cohort descriptives | | | |
| --- | --- | --- | --- |
|  | **NSHD46** | **NCDS58** | **BCS70** |
| ***Early life*** |  |  |  |
| Non-manual job by father, % (95% CI) | 40.3 (38.8-41.9) | 32.1 (31.3-34.9) | 36.7 (35.9-37.5) |
| NVQ level 4+ | 8.5 (7.7-9.4) | 24.0 (23.3-24.8) | 33,8 (32.6-35.0) |
| ***30s*** |  |  |  |
| Data collection year | 1982 | 1991 | 2003-2004 |
| N | 4,388 | 16,173 | 16,787 |
| Female, % (95% CI) | 48.5 (47.0-50.0) | 48.8 (48.0-49.6) | 48.6 (47.8-49.4) |
| Age, mean (SD) | 36^a^ | 33^a^ | 33.90 (0.01) |
| Economically inactive, % (95% CI) | 13.4 (12.3-14.4) | 8.9 (8.3-9.4) | 10.3 (9.7-11.0) |
| Psychological distress, % (95% CI) | 7.3 (6.3-8.3) | 9.3 (8.6-10.0) | 18.7 (17.7-19.7) |
| Cardiometabolic disease, % (95% CI) | 7.1 (6.1-8.2) | 9.1 (8.5-9.7) | 12.4 (11.2-13.5) |
| Back pain, % (95% CI) | 18.6 (17.2-20.1) | 29.2 (28.1-30.2) | 12.8 (12.0-13.7) |
| In relationship, % (95% CI) | 84.8 (83.5-86.2) | 84.0 (83.1-84.8) | 69.8 (68.5-71.1) |
| Own their home, % (95% CI) | 76.2 (74.5-77.8) | 73.4 (72.3-74.5) | 65.5 (63.8-67.3) |
| Children or 2+ people in the household | 82.1 (80.8-83.5) | 73.4 (72.3-74.6) | 60.2 (58.8-61.5) |
| Antidepressant use, % (95% CI) | 1.3 (0.9-1.8) | - | - |
| ***40s*** |  |  |  |
| Data collection year | 1989 | 2000 | 2012 |
| N | 4,396 | 16,090 | 16,639 |
| Female, % (95% CI) | 48.5 (47.0-50.0) | 48.9 (48.2-49.7) | 48.7 (48.0-49.5) |
| Age, mean (SD) | 43.5 (0.18) | 42^a^ | 42.02 (0.01) |
| Economically inactive, % (95% CI) | 11.1 (10.0-12.2) | 12.1 (11.5-12.8) | 13.3 (12.4-14.2) |
| Psychological distress, % (95% CI) | 18.7 (17.3-20.1) | 14.8 (13.9-15.6) | 22.6 (21.2-24.0) |
| Cardiometabolic disease, % (95% CI) | 15.9 (14.5-17.4) | 16.0 (15.1-16.8) | 16.7 (15.5-17.9) |
| Back pain, % (95% CI) | 28.7 (27.1-30.3) | 16.6 (15.8-17.5) | 19.9 (18.5-21.3) |
| In relationship, % (95% CI) | 76.8 (75.3-78.3) | 76.5 (75.5-77.6) | 56.4 (54.9-57.9) |
| Own their home, % (95% CI) | 82.4 (80.9-83.8) | 73.8 (72.7-74.9) | 62.6 (61.3-63.9) |
| Children or 2+ people in the household | 75.5 (74.0-77.0) | 71.2 (70.1-72.4) | 69.7 (68.4-71.1) |
| Antidepressant use, % (95% CI) | 2.3 (1.7-2.9) | - | - |
| *50s* |  |  |  |
| Data collection year | 1999 | 2008-2009 | 2021-2024 |
| N | 4,299 | 15,805 | 16,749 |
| Female | 48.6 (47.1-50.1) | 49.1 (48.3-49.9) | 48.8 48.1-49.6) |
| Age, mean (SD) | 53.5 (0.18) | 50.16 (0.01) | 51.96 (0.01) |
| Economically inactive, % (95% CI) | 22.0 (20.5-23.5) | 15.4 (14.7-16.1) | 14.1 (13.3-14.9) |
| ^a^Age is only available as a round number. | | | |

| Supplementary Table 3. Breakdown of economic inactivity status by work-seeking | | | |
| --- | --- | --- | --- |
|  | **NSHD46** | **NCDS58** | **BCS70** |
| ***30s*** |  |  |  |
| Economically inactive, N | 586 | 1435 | 1734 |
| % Looking for work | 50.3 | 60.7 | 40.5 |
| % Not looking for work | 49.7 | 39.3 | 59.5 |
| ***40s*** |  |  |  |
| Economically inactive, N | 485 | 1954 | 2211 |
| % Looking for work | 44.5 | 27.5 | 38.1 |
| % Not looking for work | 55.5 | 72.5 | 61.9 |
| ***50s*** |  |  |  |
| Economically inactive, N | 945 | 2433 | 2355 |
| % Looking for work | 17.2 | 26.3 | 21.3 |
| % Not looking for work | 82.8 | 73.7 | 78.7 |

| Supplementary Table 4. Interaction results | | | |
| --- | --- | --- | --- |
|  | **NSHD46** | **NCDS58** | **BCS70** |
| **30s-40s** | | | |
| **Psychological distress x back pain** | | | |
| Psychological distress | 1.39 [0.99,1.96] | 1.85 [1.51,2.27] | 2.08 [1.71,2.54] |
| Back pain | 1.03 [0.76,1.39] | 1.21 [1.05,1.40] | 1.21 [0.94,1.56] |
| Multiplicative interaction | 0.87 [0.42,1.80] | 0.92 [0.71,1.21] | 0.74 [0.48,1.15] |
| Additive interaction (RERI) | -0.15 [-1.08,0.78] | 0.00 [-0.56,0.56] | -0.43 [-1.21,0.36] |
| **Psychological distress x cardiometabolic disease** | | | |
| Psychological distress | 1.33 [0.96,1.84] | 1.88 [1.58,2.24] | 2.03 [1.68,2.45] |
| Cardiometabolic disease | 1.36 [0.91,2.04] | 1.53 [1.31,1.79] | 1.30 [1.00,1.68] |
| Multiplicative interaction | 0.87 [0.43,1.76] | 0.78 [0.60,1.01] | 0.84 [0.60,1.19] |
| Additive interaction (RERI) | -0.08 [-1.12,0.95] | -0.17 [-0.79,0.45] | -0.11 [-0.81,0.59] |
| **Cardiometabolic disease x pack pain** | | | |
| Cardiometabolic disease | 1.49 [1.01,2.21] | 1.50 [1.27,1.77] | 1.28 [1.06,1.54] |
| Back pain | 1.07 [0.81,1.41] | 1.24 [1.10,1.41] | 1.12 [0.94,1.33] |
| Multiplicative interaction | 0.69 [0.32,1.52] | 0.98 [0.76,1.27] | 0.96 [0.71,1.31] |
| Additive interaction (RERI) | 0.46 [-0.81,1.72] | 0.08 [-0.4,0.57] | -0.02 [-0.46,0.42] |
| **30s-50s** | | | |
| **Psychological distress x back pain** | | | |
| Psychological distress | 1.67 [1.35,2.07] | 1.71 [1.38,2.13] | 1.98 [1.69,2.31] |
| Back pain | 1.44 [1.21,1.72] | 1.25 [1.08,1.46] | 1.39 [1.03,1.88] |
| Multiplicative interaction | 0.64 [0.42,0.98] | 0.97 [0.71,1.33] | 0.73 [0.46,1.15] |
| Additive interaction (RERI) | -0.55 [-1.21,0.10] | 0.12 [-0.47,0.71] | -0.36 [-1.21,0.48] |
| **Psychological distress x cardiometabolic disease** | | | |
| Psychological distress | 1.42 [1.12,1.81] | 1.72 [1.45,2.05] | 1.94 [1.65,2.29] |
| Cardiometabolic disease | 1.72 [1.38,2.14] | 1.63 [1.41,1.88] | 1.42 [1.13,1.77] |
| Multiplicative interaction | 1.02 [0.65,1.59] | 0.90 [0.70,1.17] | 0.80 [0.55,1.16] |
| Additive interaction (RERI) | 0.35 [-0.55,1.24] | 0.18 [-0.45,0.82] | -0.16 [-0.92,0.61] |
| **Cardiometabolic disease x pack pain** | | | |
| Cardiometabolic disease | 1.85 [1.50,2.28] | 1.64 [1.37,1.97] | 1.37 [1.11,1.69] |
| Back pain | 1.38 [1.17,1.64] | 1.29 [1.12,1.47] | 1.30 [1.04,1.61] |
| Multiplicative interaction | 0.83 [0.54,1.29] | 0.99 [0.79,1.26] | 0.91 [0.63,1.31] |
| Additive interaction (RERI) | 0.19 [-0.11,1.72] | 0.17 [-0.29,0.63] | -0.06 [-0.62,0.50] |
| **40s-50s** | | | |
| **Psychological distress x back pain** | | | |
| Psychological distress | 1.21 [1.00, 1.47] | 1.68 [1.47,1.93] | 1.44 [1.21,1.72] |
| Back pain | 1.18 [0.99,1.41] | 1.31 [1.14,1.51] | 1.20 [0.98,1.46] |
| Multiplicative interaction | 1.17 [0.87,1.57] | 0.95 [0.78,1.17] | 1.26 [0.94,1.67] |
| Additive interaction (RERI) | 0.28 [-0.10,0.66] | 0.11 [-0.32,0.55] | 0.53 [-0.01,1.06] |

*Table continued overleaf*

*Table continued*

| **Psychological distress x cardiometabolic disease** | | | |
| --- | --- | --- | --- |
| Psychological distress | 1.31 [1.11, 1.54] | 1.78 [1.55,2.03] | 1.50 [1.27,1.79] |
| Cardiometabolic disease | 1.43 [1.16,1.78] | 1.46 [1.24,1.71] | 1.14 [0.92,1.43] |
| Multiplicative interaction | 0.94 [0.69,1.29] | 0.84 [0.70,1.02] | 1.18 [0.89,1.57] |
| Additive interaction (RERI) | 0.02 [-0.45,0.49] | -0.05 [-0.50,0.40] | 0.39 [-0.11,0.88] |
| **Cardiometabolic disease x pack pain** | | | |
| Cardiometabolic disease | 1.57 [1.30,1.90] | 1.43 [1.23,1.65] | 1.27 [1.06,1.52] |
| Back pain | 1.36 [1.17,1.58] | 1.40 [1.24,1.58] | 1.38 [1.21,1.58] |
| Multiplicative interaction | 0.75 [0.53,1.07] | 0.94 [0.77,1.15] | 1.08 [0.85,1.36] |
| Additive interaction (RERI) | 0.25 [-0.01,0.51] | 0.04 [-0.36,0.45] | 0.24 [-0.22,0.69] |
| Age pairings represent age at health assessment and economic activity assessment. Statistics derived from fully adjusted models. RR: risk ratio, CI: confidence intervals. | | | |
